# COVID-19 risk perception and vaccine acceptance in individuals with chronic disease

**DOI:** 10.1101/2021.03.17.21253760

**Authors:** Brianna A. Smith, Emily E. Ricotta, Jennifer L. Kwan, Nicholas G. Evans

## Abstract

**Background:** COVID-19 disproportionately affects those with preexisting conditions, but little research has determined whether those with chronic diseases view the pandemic itself differently - and whether there are differences between chronic diseases. We theorized that while individuals with respiratory disease or autoimmune disorders would perceive greater threat from COVID-19 and be more supportive of non-pharmaceutical interventions (NPIs), those with autoimmune disorders would be less likely to support vaccination-based interventions.

**Methods:** We conducted a two-wave online survey conducted in February and November 2021 asking respondents their beliefs about COVID-19 risk perception, adoption and support of interventions, willingness to be vaccinated against COVID-19, and reasons for vaccination. Regression analysis was conducted to assess the relationship of respondents reporting a chronic disease and COVID-19 behaviors and attitudes, compared to healthy respondents adjusting for demographic and political factors.

**Results:** In the initial survey, individuals reporting a chronic disease had stronger both stronger feelings of risk from COVID-19 as well as preferences for NPIs than healthy controls. The only NPI that was still practiced significantly more compared to healthy controls in the resample was limiting trips outside of the home. Support for community-level NPIs was higher among individuals reporting a chronic disease than healthy controls and remained high among those with respiratory diseases in sample 2. Vaccine acceptance produced more divergent results: those reporting chronic respiratory diseases were 6% more willing to be vaccinated than healthy controls, while we found no significant difference between individuals with autoimmune diseases and healthy controls. Respondents with chronic respiratory disease and those with autoimmune diseases were more likely to want to be vaccinated to protect themselves from COVID-19, and those with an autoimmune disease were more likely to report fear of a bad vaccine reaction as the reason for vaccine hesitancy. In the resample, neither those with respiratory diseases nor autoimmune diseases reported being more willing to receive a booster vaccine than healthy controls.

**Conclusions:** It is not enough to recognize the importance of health in determining attitudes: nuanced differences between conditions must also be recognized.

## Background

The COVID-19 pandemic disproportionately affects individuals with comorbidities (1–4), but communication and risk perception around patient groups remains understudied. Recent work outside the US has demonstrated that individuals with comorbidities are significantly less likely to refuse vaccination and are more likely to take personal health-protective measures against COVID-19 (5,6). US studies show broad vaccine acceptance for those with underlying medical conditions (7), but global reviews have noted conflicting results between chronic disease status and vaccine acceptance (8). This signals a gap in understanding key chronic illness patient groups’ risk perceptions of COVID-19, their beliefs about personal and community-level nonpharmaceutical interventions (NPIs), and willingness to be vaccinated.

Understanding acceptance of NPIs and vaccination is critical to ongoing response efforts, especially as uptake of vaccines and boosters has stalled. In earlier phases of the pandemic concerns were raised about the risk of developing severe COVID-19 for individuals with chronic respiratory and autoimmune diseases. The latter’s potential contraindication for vaccine candidates was also of concern, and these individuals remain under-addressed in CDC vaccine guidance due to lack of data (9). These uncertainties disproportionately affect marginalized groups: non-Hispanic blacks are more likely to experience complications and death from chronic respiratory diseases than non-Hispanic whites (10) but are less widely vaccinated against COVID-19 (11), while women are more likely to have autoimmune diseases than men (12). As federal and state authorities abandoned NPIs in favor of vaccine-only mitigation (13), patients with chronic illnesses may have altered risk attitudes if they felt their best options for collective protection had been abandoned by the public.

Our study identifies the relationship between an individual’s self-identified chronic illness, how they perceive COVID-19 risks, engage with individual and community-level NPIs, and weigh the benefits and risks of vaccination in the US context. Self-identification is important as these categories do not strictly track medical diagnoses, but rather the relationship between medical indications, physical or cognitive capacities, and social function (14). For example, merely having a chronic impairment of respiratory function may not necessarily constitute identification as chronically ill, unless it compromises an individual’s ability to achieve their goals or social aims: exercise-induced asthma may not be identified as a chronic illness if it is simply an inconvenience easily managed with an inhaler, versus severe asthma that is unable to be easily managed or excludes an individual from social settings on air quality warning days, periods of high airborne allergen concentrations, and so on.

Our follow-up survey, conducted in the same population, assessed participants’ fatigue with the pandemic, their continued willingness to undertake individual and community-level NPIs, vaccine enthusiasm, and the persistence of these beliefs from pre- to post-COVID-19 vaccine availability. We hypothesized that those with chronic respiratory and autoimmune disease would be both more concerned about COVID-19 and more likely to support risk-mitigation than healthy individuals, while those with autoimmune disease would be less likely to vaccinate due to ambiguous and changing CDC guidance.

## Methods

### Survey

Participants were contacted through Prolific’s survey platform, which recruits a large and diverse pool of potential participants through social media, physical flyers, and referrals. While this is a non-probability convenience sample, it is more diverse than most convenience samples and researchers have successfully replicated established studies through the platform (15,16). Unlike random sample, Prolific allowed oversampling of respondents who had previously self-reported chronic respiratory disease or autoimmune disease (3,124). US residents 18 years of age or older participated in the study in February 2021. Of those, we included 2,535 individuals in this analysis: 478 who reported having any autoimmune disorder; 618 who reported having asthma, chronic obstructive pulmonary disease, or any other chronic respiratory condition; 136 who reported having both; and 1303 who reported no chronic condition (“healthy controls”). 589 respondents reported some other chronic illness (such as cancer or diabetes) and were excluded from analysis as we did not oversample these categories and did not have a large enough sample to accurately estimate an effect for any other illness category. A resample was conducted in November 2021: 55% of initial respondents participated, including 54% of those with respiratory diseases, 61% of those with autoimmune disorders, and 57% of those with both. Balance tests comparing the demographics of survey 2 participants to those whom we were unable to recontact from survey 1 are available in the appendix. Women, older adults, non-Hispanic whites, non-students, Republicans, and those with autoimmune disease were somewhat more likely to participate in survey 2. These factors are controlled for in the following analyses, in addition to further demographic controls described below.

In both samples, participants were asked about their beliefs about the risk of COVID-19 to their health, to the public’s health, and whether the risk of COVID-19 is overblown. These risk perceptions were collected on a five-point scale ranging from “strongly agree” to “strongly disagree,” and then normalized to a [0,1] interval where 1 indicates strong agreement. The full survey is available in the Supplement. In the resample, we additionally assessed ‘pandemic fatigue.’ First, questions based on Johansson et al.’s mental fatigue scale (17) assessed the extent to which participants felt stressed, irritable, or unable to cope with the COVID-19 pandemic. Participants answered on a five-point agree/disagree scale, normalized to a [0,1] interval where 1 indicates the highest level of fatigue. Responses to the three questions were averaged to create an emotional fatigue scale (α = 0.76). An additional question assessed the extent to which participants were ‘over’ the pandemic and were ready to go back to normal. Again, participants answered on a five-point scale normalized to a [0,1] interval.

Adoption of individual risk mitigation measures were collected on a six-point true/false scale normalized to a [0,1] interval where 1 indicates greatest adoption of that measure. Individual risk mitigation measures were reducing trips; mask wear; working from home; handwashing; and maintaining physical distance. In the resample, participants were asked about the same set of individual risk mitigation measures, excluding working from home. Approval of community-level NPIs (mask mandates, limits on indoor dining, limits on in-person worship services, lockdown of non-essential travel, and school closures) were measured on a six-point approve/disapprove scale, normalized to a [0,1] interval where 1 indicates strong approval. In the resample, participants evaluated the same set of policies, with the addition of two questions about a vaccine mandate and vaccine ‘passports’.

In the first sample, respondents indicated whether they were willing, unwilling, or had already received a COVID-19 vaccine on a seven-point scale, then provided reasons for their vaccine decision. For those willing to be or already vaccinated, reasons included protecting themselves, protecting others, belief the vaccine had been fully tested, the vaccine was safe, a desire to get back to normal, or a need to be vaccinated for work or other activities. For those unwilling to get a vaccine, potential reasons included belief that COVID-19 is not a serious health threat, concern about a bad reaction to the vaccine, belief the vaccine had not been fully tested, the vaccine was not safe, or opposition to vaccines in general. Participants could select multiple reasons or write their own. Willingness to receive the vaccine was normalized to a [0,1] interval where 1 indicates either that participants would “definitely get the vaccine” or had already been vaccinated. In the resample, participants were asked if they were vaccinated, unvaccinated, or partially vaccinated. They then indicated whether they were willing, unwilling, or had already received a COVID-19 vaccine booster on a seven-point scale.

Age, race/ethnicity, sex, household income, educational level, geographic region, rural or urban residence, partisanship, employment, and education demographics were collected for the first sample (Table 1). These roughly matched 2019 American Community Survey (ACS) demographics, with the exception that participants were somewhat more likely to be white than the national average, and specific demographics varied by illness category. Respondents were also asked in both samples if they had been diagnosed with COVID-19 in the past, and whether a family member or close friend had been diagnosed with COVID-19.

**Table 1.** Characteristics of the initial sample respondents by reported illness.

| Variable | Self-reported disease state |  |  |  |  |
| --- | --- | --- | --- | --- | --- |
|  | None | Respiratory | Autoimmune | Both | US |
|  | (N=1303) | (N=618) | (N=478) | (N=136) | (2019 ACS) |
| **Age - average years [SD]** | 32.6 [11.3] | 35.8 [14.2] | 38.5 [12.6] | 39.2 [14.5] | 38.4 (median) |
| **Race – No. (%)** |  |  |  |  |  |
| **Non-Hispanic White** | 872 (67.7) | 428 (70.2) | 381 (80.4) | 108 (79.4) | 60.10% |
| **Black** | 82 (6.4) | 45 (7.38) | 22 (4.6) | 6 (4.4) | 12.20% |
| **Hispanic or Latino** | 63 (4.9) | 25 (4.1) | 22 (4.6) | 4 (2.9) | 18.50% |
| **Asian American** | 180 (14.0) | 58 (9.5) | 21 (4.4) | 7 (5.2) | 5.60% |
| **Native American** | 6 (0.5) | 6 (0.98) | 1 (0.2) | 2 (1.5) | 0.70% |
| **Two or more Ethnicities** | 86 (6.7) | 48 (7.9) | 27 (5.7) | 9 (6.6) | 2.80% |
| **Female – No. (%)** | 673 (51.8) | 359 (58.1) | 361 (75.5) | 111 (81.6) | 50.80% |
| **Education - median** | 4-year College Degree | 2-year College Degree | 4-year College Degree | 2-year College Degree | 4-year College Degree |
| **Household Income – Median** | \$60,000-69,999 | \$50,000-59,999 | \$50,000-59,999 | \$40,000-49,999 | \$68,703 |
| **Region – No. (%)** |  |  |  |  |  |
| **West** | 337 (26.1) | 157 (25.6) | 105 (22.1) | 30 (22.1) | 23.90% |
| **Midwest** | 258 (20.0) | 132 (21.5) | 93 (19.5) | 32 (23.5) | 20.80% |
| **South** | 448 (34.7) | 199 (32.5) | 182 (38.2) | 47 (34.6) | 38.30% |
| **Northeast** | 247 (19.2) | 125 (20.4) | 96 (20.2) | 27 (19.9) | 17.10% |
| **Urban-Rural – mean (0 = Urban; 1 = Rural)** | 0.42 | 0.44 | 0.47 | 0.47 | — |
| **Partisanship – mean (0 = Strong Democrat; 1 = Strong Republican)** | 0.3 | 0.26 | 0.28 | 0.32 | — |
| **Had COVID themselves – No. (%)** | 88 (6.8) | 39 (6.3) | 35 (7.3) | 16 (11.8) | 7% (Ref. 35) |
| **Family/close friends had COVID – No. (%)** | 694 (53.3) | 339 (54.9) | 297 (62.1) | 82 (60.3) | 67% (Ref. 36) |
| **Students – No. (%)** | 341 (27.1) | 157 (25.8) | 91 (19.2) | 30 (22.4) | 8.40% |
| **Employed full time – No. (%)** | 569 (48.5) | 212 (38.7) | 194 (43.5) | 23 (19.3) | 59.50% |

### Data analysis

We conducted regression analyses to evaluate the association between disease status and COVID-19 behaviors or attitudes, as well as the change in attitudes by disease from survey 1 to survey 2 using R version 4.0.2 and STATA version 17 (18), controlling for the demographics listed above. For risk perceptions, pandemic fatigue, individual risk mitigation behaviors, approval of community-level NPIs, and vaccine/booster willingness we conducted OLS regressions. For vaccine attitudes, we conducted logistic regression. To test whether the slopes of the regression coefficients for “COVID-19 is a threat to me” and “COVID-19 is a threat to the public” were significantly different from one another we used seemingly unrelated regression to account for possible correlation of the equation errors (using the “systemfit” package) (19) and tested the linear hypothesis using an asymptotic Chi-square test (“car” package) (20). To assess the effect of chronic disease status on change in outcome from survey 1 to survey 2, we conducted OLS regression controlling for survey 1 response, disease status, and the factors above to predict survey 2 response among participants who completed both surveys. Results are presented as linear regression coefficients or odds ratios and 95% confidence intervals, and predicted values and standard error for the change in response between surveys.

## Results

Regression coefficients and odds ratios can be seen in Table 2. All predictor and outcome variables except age were normalized to [0,1], so OLS regression coefficients may be interpreted as the percent change in the outcome due to the predictor variable. *Personal beliefs about COVID-19*

**Table 2.** Regression coefficients or odds ratios evaluating the association of attitudes about COVID-19 risk and willingness to vaccinate among individuals with autoimmune and respiratory diseases.

|  | Respiratory |  | Autoimmune |  |
| --- | --- | --- | --- | --- |
|  | Sample 1 | Sample 2 | Sample 1 | Sample 2 |
| Outcome | Coefficient (95% CI) | Coefficient (95% CI) | Coefficient (95% CI) | Coefficient (95% CI) |
| ***Attitudes about COVID-19*** |  |  |  |  |
| COVID-19 is a serious threat to my health | **0.12 (0.10, 0.15)** | **0.15 (0.11, 0.19)** | **0.11 (0.08, 0.14)** | **0.12 (0.08, 0.17)** |
| COVID-19 is a serious threat to the public's health | **0.04 (0.02, 0.06)** | **0.05 (0.01, 0.08)** | **0.03 (0.01, 0.06)** | 0.02 (-0.01, 0.06) |
| The threat of COVID-19 is overblown | **-0.06 (-0.09, -0.03)** | -0.04 (-0.08, 0.00) | **-0.04 (-0.07, -0.01)** | -0.03 (-0.08, 0.01) |
| Emotional/Mental fatigue | — | **0.06 (0.03, 0.10)** | — | **0.07 (0.03, 0.11)** |
| Need to get back to 'normal' | — | -0.04 (-0.09, 0.00) | — | -0.05 (-0.10, 0.00) |
| ***Individual-level interventions*** | — | — | — | — |
| Handwashing | 0.02 (-0.01, 0.04) | 0.02 (-0.02, 0.06) | **0.05 (0.02, 0.08)** | 0.03 (-0.01, 0.07) |
| Limiting trips outside the home | **0.03 (0.01, 0.06)** | **0.06 (0.01, 0.10)** | **0.04 (0.01, 0.07)** | **0.07 (0.02, 0.12)** |
| Mask wearing | **0.03 (0.01, 0.04)** | 0.04 (0.00, 0.08) | **0.02 (0.01, 0.04)** | 0.03 (-0.02, 0.07) |
| Social distancing | **0.03 (0.01, 0.05)** | 0.04 (0.00, 0.08) | **0.04 (0.02, 0.07)** | 0.04 (0.00, 0.08) |
| Working from home | 0.02 (-0.02, 0.07) | — | 0.03 (-0.02, 0.08) | — |
| Willingness to receive a COVID-19 vaccine* | **0.06 (0.03, 0.09)** | — | 0.02 (-0.01, 0.05) | — |
| Willingness to receive a COVID-19 booster** | — | 0.02 (-0.02, 0.06) | — | -0.03 (-0.07, 0.01) |
| ***Community-level interventions*** | — | — | — | — |
| Limits on indoor dining | **0.06 (0.03, 0.08)** | 0.03 (-0.02, 0.07) | 0.03 (0.00, 0.06) | 0.01 (-0.04, 0.05) |
| Limits on in-person worship services | **0.04 (0.01, 0.07)** | **0.06 (0.02, 0.11)** | 0.03 (-0.01, 0.06) | 0.03 (-0.02, 0.08) |
| Lockdown of all non-essential travel | **0.05 (0.01, 0.08)** | 0.04 (-0.01, 0.09) | **0.06 (0.02, 0.09)** | 0.05 (0.00, 0.10) |
| Mandatory mask-wearing in public | **0.05 (0.02, 0.07)** | **0.05 (0.01, 0.09)** | **0.03 (0.01, 0.06)** | 0.02 (-0.03, 0.06) |
| School closures | **0.05 (0.01, 0.08)** | **0.07 (0.02, 0.11)** | **0.04 (0.01, 0.07)** | 0.05 (0.00, 0.10) |
| Vaccine mandates | — | **0.07 (0.02, 0.12)** | — | 0.04 (-0.01, 0.09) |
| Vaccine passports | — | **0.07 (0.02, 0.11)** | — | -0.01 (-0.05, 0.05) |
| ***Vaccine attitudes*** | **Odds ratio (95% CI)** | — | **Odds ratio (95% CI)** | — |
| Willing: Want to protect myself from COVID-19 | **1.88 (1.30, 2.70)** | — | **1.50 (1.01, 2.23)** | — |
| Willing: Want to protect others from COVID-19 | 1.16 (0.85, 1.58) | — | 1.07 (0.76, 1.52) | — |
| Willing: Vaccine is safe | 1.21 (0.93, 1.56) | — | 1.13 (0.85, 1.51) | — |
| Willing: Vaccine has been fully tested | 1.08 (0.84, 1.38) | — | 1.07 (0.81, 1.42) | — |
| Willing: Want to return to work | **1.49 (1.14, 1.95)** | — | 1.18 (0.86, 1.61) | — |
| Willing: Want life to return to normal | 1.07 (0.82, 1.40) | — | 0.90 (0.67, 1.21) | — |
| Unwilling: Don't believe in vaccines | 0.96 (0.36, 2.62) | — | 0.80 (0.28, 2.32) | — |
| Unwilling: Afraid of bad reaction | 1.91 (0.99, 3.66) | — | **2.47 (1.29, 4.72)** | — |
| Unwilling: Vaccine is not safe | 1.55 (0.83, 2.91) | — | 1.01 (0.53, 1.89) | — |
| Unwilling: Vaccine has not been fully tested | 1.31 (0.66, 2.59) | — | 0.85 (0.44, 1.65) | — |
| Unwilling: COVID-19 is not a serious threat | 0.58 (0.23, 1.44) | — | 0.37 (0.14, 0.96) | — |
| Fully vaccinated | — | **1.85 (1.18, 2.92)** | — | 0.81 (0.53, 1.26) |
| *Inclusive of individuals already vaccinated. A separate regression excluding vaccinated respondents did not change the results. |  |  |  |  |
| **Inclusive of individuals already boosted. In a separate regression excluding boosted respondents, participants with autoimmune disorders were somewhat less likely to be willing to receive a booster; the results are available in the appendix. |  |  |  |  |

In the initial sample, respondents reporting chronic respiratory or autoimmune diseases were significantly more likely than healthy controls to report that COVID-19 was a threat to themselves (Respiratory (B_R_) = 0.12, 95% confidence interval (CI) = 0.10 – 0.15; Autoimmune (B_A_) = 0.11, CI = 0.08 – 0.14) and to a lesser extent the public’s health (B_R_ A = 0.04, CI = 0.02 – 0.06; B_A_ = 0.03, CI = 0.01 – 0.06). In both cases, the effect of disease status on perception of threat to the respondent was significantly higher than the effect on perception of threat to the public (χ_R_^2^=21.3, p<0.001; χ_A_^2^ 2=15.2, p<0.001). Respondents reporting chronic respiratory or autoimmune diseases were also less likely than healthy controls to think the threat of COVID-19 was overblown (B_R_ = -0.06, CI = -0.09 – -0.03; B_A_ = -0.4, CI = -0.07 – -0.01). In the resample, while both groups still thought that COVID-19 was a threat to themselves (B_R_ = 0.15, CI = 0.11 – 0.19; B_A_ = 0.12, CI = 0.08 – 0.17), those with autoimmune disease were no longer more likely to believe it was a threat to the public’s health, and neither group’s greater beliefs about the threat of COVID-19 being overblown persisted. Both chronic condition groups felt more emotional/mental fatigue than healthy controls (B_R_ = 0.06, CI = 0.03 – 0.10; B_A_ = 0.07, CI = 0.03 – 0.11), but neither felt more need to get back to “normal”.

### Acceptance of NPIs

Acceptance of NPIs in the first sample was broadly concordant with COVID-19 risk perceptions: individuals reporting a chronic disease had stronger preferences for NPIs. They were more likely to wear masks outside the home (B_R_ = 0.03, CI = 0.01 – 0.04; B_A_ = 0.02, CI = 0.01 – 0.04), physically distance (B_R_ = 0.03, CI = 0.01 – 0.05; B_A_ = 0.04, CI = 0.02 – 0.07), and decrease trips outside the home (B_R_ = 0.03, CI = 0.01 – 0.06; B_A_ = 0.04, CI = 0.01 – 0.07). Interestingly, the only NPI that was still practiced significantly more compared to healthy controls in the resample was limiting trips outside of the home (B_R_ = 0.06, CI = 0.01 – 0.10; B_A_ = 0.07, CI = 0.02 – 0.12).

Support for community-level NPIs was higher among individuals reporting a chronic disease than healthy controls and remained high among those with respiratory diseases in sample 2. In sample 1, both groups were more likely to support prohibitions on indoor dining (B_R_ = 0.06, CI = 0.03 – 0.08; B_A_ = 0.03, CI = 0.002 – 0.06), broad lockdowns (B_R_ = 0.05, CI = 0.01 – 0.08; B_A_ = 0.06, CI = 0.02 – 0.09), mask mandates (B_R_ = 0.05, CI = 0.02 – 0.07; B_A_ = 0.03, CI = 0.01 – 0.06), and school closures (B_R_ = 0.05, CI = 0.01 – 0.08; B_A_ = 0.04, CI = 0.01 – 0.07). Only limits on in-person worship services diverged: while respondents reporting chronic respiratory diseases were significantly more likely than controls to support limits (B_R_ *=* 0.04, CI = 0.01 – 0.07), individuals reporting autoimmune diseases were no more or less likely to prefer those limits than healthy controls (B_A_ *=* 0.03, CI = -0.01 – 0.06). In the resample, those with respiratory diseases still supported limits on in-person worship services, mandatory mask wearing in public, and school closures, and also supported vaccine mandates (B_R_ *=* 0.07, CI = 0.02 – 0.12) and passports (B_R_ *=* 0.07, CI = 0.02 – 0.11), but those with autoimmune diseases showed no difference in support compared to healthy controls.

### Vaccine acceptance

Vaccine acceptance produced more divergent results. Respondents who reported chronic respiratory diseases were 6% more willing to be vaccinated than healthy controls (CI = 0.03 – 0.09), while we found no significant difference between individuals with autoimmune diseases and healthy controls (B_A_ = 0.02, CI = -0.01 – 0.05). When assessing reasons for being willing or having been vaccinated, respondents with chronic respiratory disease and those with autoimmune diseases were more likely to want to be vaccinated to protect themselves from COVID-19 (OR_R_ = 1.88, CI = 1.30 – 2.70; OR_A_ = 1.50, CI = 1.01 – 2.23). Respondents reporting a chronic respiratory disease were also more likely to want to safely return to work (OR_R_ = 1.49, CI = 1.14 – 1.95). Individuals with autoimmune diseases were the only group to have a significant association with a particular cause for vaccine hesitancy: they were more likely to report fear of a bad vaccine reaction as the reason for unwillingness (OR_A_ = 2.47; CI = 1.29 – 4.72). Respondents with autoimmune diseases were also less likely to say that their unwillingness was due to not seeing COVID-19 as a threat (OR_A_ = 0.37; CI = 0.14 – 0.96). In the resample, neither those with respiratory diseases nor autoimmune diseases reported being more willing to receive a booster vaccine than healthy controls.

Acceptance of certain behaviors within disease groups did change from sample 1 to sample 2, but disease status *itself* was rarely responsible for changing attitudes and behaviors (Table 3). For example, support for limits on indoor dining decreased from sample 1 to sample 2 regardless of disease status. The exceptions are that those with respiratory diseases became more likely to get the vaccine even controlling for sample 1 vaccine intention, and those with autoimmune disorders became more likely to avoid trips outside the home even controlling for sample 1 behavior. In contrast, partisanship, for example, was significantly and consistently associated with changes in attitudes towards COVID-19 mitigation measures over time (see supplement).

**Table 3.**
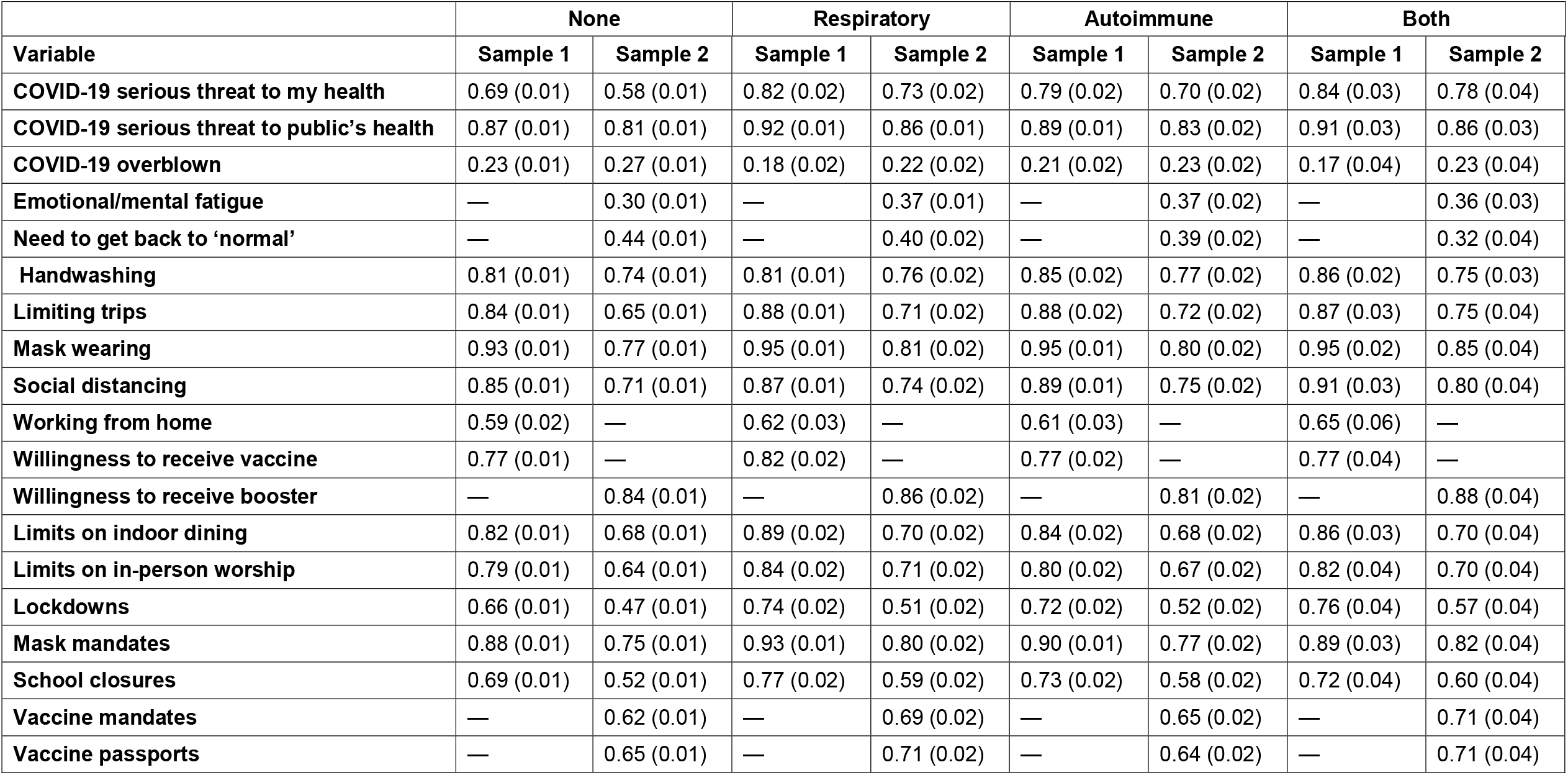
Change in COVID-19 Attitudes, sample 1 to sample 2. Predicted levels of agreement with standard errors in parentheses. Only participants who participated in both samples are included.

|  | None |  | Respiratory |  | Autoimmune |  | Both |  |
| --- | --- | --- | --- | --- | --- | --- | --- | --- |
| Variable | Sample 1 | Sample 2 | Sample 1 | Sample 2 | Sample 1 | Sample 2 | Sample 1 | Sample 2 |
| **COVID-19 serious threat to my health** | 0.69 (0.01) | 0.58 (0.01) | 0.82 (0.02) | 0.73 (0.02) | 0.79 (0.02) | 0.70 (0.02) | 0.84 (0.03) | 0.78 (0.04) |
| **COVID-19 serious threat to public's health** | 0.87 (0.01) | 0.81 (0.01) | 0.92 (0.01) | 0.86 (0.01) | 0.89 (0.01) | 0.83 (0.02) | 0.91 (0.03) | 0.86 (0.03) |
| **COVID-19 overblown** | 0.23 (0.01) | 0.27 (0.01) | 0.18 (0.02) | 0.22 (0.02) | 0.21 (0.02) | 0.23 (0.02) | 0.17 (0.04) | 0.23 (0.04) |
| **Emotional/mental fatigue** | — | 0.30 (0.01) | — | 0.37 (0.01) | — | 0.37 (0.02) | — | 0.36 (0.03) |
| **Need to get back to 'normal'** | — | 0.44 (0.01) | — | 0.40 (0.02) | — | 0.39 (0.02) | — | 0.32 (0.04) |
| **Handwashing** | 0.81 (0.01) | 0.74 (0.01) | 0.81 (0.01) | 0.76 (0.02) | 0.85 (0.02) | 0.77 (0.02) | 0.86 (0.02) | 0.75 (0.03) |
| **Limiting trips** | 0.84 (0.01) | 0.65 (0.01) | 0.88 (0.01) | 0.71 (0.02) | 0.88 (0.02) | 0.72 (0.02) | 0.87 (0.03) | 0.75 (0.04) |
| **Mask wearing** | 0.93 (0.01) | 0.77 (0.01) | 0.95 (0.01) | 0.81 (0.02) | 0.95 (0.01) | 0.80 (0.02) | 0.95 (0.02) | 0.85 (0.04) |
| **Social distancing** | 0.85 (0.01) | 0.71 (0.01) | 0.87 (0.01) | 0.74 (0.02) | 0.89 (0.01) | 0.75 (0.02) | 0.91 (0.03) | 0.80 (0.04) |
| **Working from home** | 0.59 (0.02) | — | 0.62 (0.03) | — | 0.61 (0.03) | — | 0.65 (0.06) | — |
| **Willingness to receive vaccine** | 0.77 (0.01) | — | 0.82 (0.02) | — | 0.77 (0.02) | — | 0.77 (0.04) | — |
| **Willingness to receive booster** | — | 0.84 (0.01) | — | 0.86 (0.02) | — | 0.81 (0.02) | — | 0.88 (0.04) |
| **Limits on indoor dining** | 0.82 (0.01) | 0.68 (0.01) | 0.89 (0.02) | 0.70 (0.02) | 0.84 (0.02) | 0.68 (0.02) | 0.86 (0.03) | 0.70 (0.04) |
| **Limits on in-person worship** | 0.79 (0.01) | 0.64 (0.01) | 0.84 (0.02) | 0.71 (0.02) | 0.80 (0.02) | 0.67 (0.02) | 0.82 (0.04) | 0.70 (0.04) |
| **Lockdowns** | 0.66 (0.01) | 0.47 (0.01) | 0.74 (0.02) | 0.51 (0.02) | 0.72 (0.02) | 0.52 (0.02) | 0.76 (0.04) | 0.57 (0.04) |
| **Mask mandates** | 0.88 (0.01) | 0.75 (0.01) | 0.93 (0.01) | 0.80 (0.02) | 0.90 (0.01) | 0.77 (0.02) | 0.89 (0.03) | 0.82 (0.04) |
| **School closures** | 0.69 (0.01) | 0.52 (0.01) | 0.77 (0.02) | 0.59 (0.02) | 0.73 (0.02) | 0.58 (0.02) | 0.72 (0.04) | 0.60 (0.04) |
| **Vaccine mandates** | — | 0.62 (0.01) | — | 0.69 (0.02) | — | 0.65 (0.02) | — | 0.71 (0.04) |
| **Vaccine passports** | — | 0.65 (0.01) | — | 0.71 (0.02) | — | 0.64 (0.02) | — | 0.71 (0.04) |

## Discussion

Over two nationwide surveys, we found individuals with self-reported chronic respiratory or autoimmune conditions were significantly more likely to be concerned about COVID-19’s threat to the public and, to a significantly greater extent, more concerned about their personal threat from COVID-19 compared to respondents without a chronic illness. This highlights the significant internalization of risk messaging in these communities which could provide a basis for choosing strategies to communicate public health information based on self or community interests.

### Chronic illness, risk perception, and NPI uptake

Compared to studies evaluating individuals with medically confirmed chronic illnesses (21), our respondents were enrolled based on self-identified disease. While possibly permitting misclassification, this methodology provides information about risk perception based on an individual’s beliefs about their disease state, rather than their diagnosis. Especially for diseases that are difficult to diagnose such as autoimmune diseases, self-identification may be a better proxy for risk attitudes (and subsequent uptake of interventions) than medical diagnosis alone. Indeed, we find that respondents with self-reported comorbidities potentially associated with worse COVID-19 clinical outcomes have significantly greater willingness to adopt individual risk management behaviors, and to support a variety of community-level interventions.

The relationship between chronic illness and identity may be important as part of policies directed at changing health behaviors. Chronic illness and disability, when viewed as a medical indication, may diverge from an individual’s experience of barriers to social participation, membership in a community, or as part of collective action to achieve policy aims (22). Appealing to individuals’ self-identified health state may be useful in promoting individual and community public health interventions precisely because it signals membership in a group that may be marginalized in a health crisis, and thus a need for action. For example, one in twelve Americans has asthma (23); framing risk messaging in terms of their comorbidity may generate more support for individual and community level public health measures than more general messaging. A targeted appeal to these individuals may also produce network benefits: given the high prevalence of chronic diseases, most people are likely to know at least one person whose risk perceptions match our findings and may be sympathetic to their concerns.

### Vaccine acceptance

Individuals reporting chronic respiratory diseases, but not autoimmune diseases, were more willing to receive their primary vaccine series than controls. While few vaccine uptake studies have focused on individuals with respiratory diseases, one study found that individuals with medically confirmed autoimmune diseases were equally willing to be vaccinated as healthy controls (21).

Among those willing or already vaccinated in the initial survey, both chronic disease groups reported wanting to protect themselves as a motivating factor for vaccination, confirming previous COVID-19 vaccine attitude studies (21,24–26). Additionally, respondents with chronic respiratory diseases but not autoimmune diseases were significantly more likely to want to do so because it would help them get back to work. Respondents with autoimmune diseases were significantly more likely to be vaccine hesitant due to concerns about adverse reactions than controls; this was not the case for individuals who reported chronic respiratory diseases. This relationship may reflect concerns that mRNA vaccines currently dominating the US market may cause higher rates of adverse events in individuals being treated for autoimmune diseases (7,21,24,23). This reflects previous vaccine hesitancy due to fear of side effects cited as one of the most prevalent barriers to vaccine uptake during the 2009 H1N1 influenza pandemic (28-31).

There was no difference in willingness to get a booster between either disease group and healthy controls. Recent work has argued that elevated risks among individuals with chronic illnesses will likely affect their booster vaccine intentions (32), but we found no effect. It may be that the difference was in part due to the time of sampling—the first study was conducted in May 2021 at the height of vaccination, where our resample was in November 2021, when vaccination rates had plateaued and public messaging for boosting was just beginning to reach the mainstream.

The heterogeneity in key aspects of health behaviors, most significantly in vaccination, is an important finding for individual and public health communication. If specific patient populations are more likely to be vaccine hesitant than others, medical specialty groups could focus on patient engagement for vaccine uptake earlier in a pandemic. The Kaiser Family Foundation has noted that for those Americans who are still “wait and see” over the decision to be vaccinated (33), the effectiveness of vaccination to reduce death and hospitalization is most likely to encourage individuals to change their minds. Given the safety concerns and internalized COVID-19 risk among individuals with autoimmune diseases who were unwilling to be vaccinated, specialists with a higher volume of autoimmune patients are an important resource for helping individuals navigate the decision to be vaccinated against pandemic diseases. While we did not examine whether vaccination motivations were influenced by physicians or other sources, it is critical to understand where information about vaccine safety and efficacy is provided in general, and specifically to vulnerable populations.

### Changes in attitudes over time

Over time, these trends were maintained or became more pronounced. Those with chronic illnesses were more fatigued by the pandemic, but less likely to believe it was time to get back to normal. Thus, while pandemic fatigue is defined by the WHO as “express[ing] itself as emerging demotivation to engage in protective behaviors and seek COVID-19-related information and as complacency, alienation and hopelessness,” (34) our findings show that demotivation and alienation/hopelessness may come apart for individuals whose risk perceptions may push them to endorse and pursue mitigation strategies even as they experience greater emotional and mental fatigue than the rest of the population.

Both chronic illness groups remained more likely to believe COVID-19 was a threat to their personal health, but diverged in what they believed should be done about it. Those with respiratory disease viewed COVID-19 more as a general threat, to be controlled with vaccination, masking, and limits on gatherings. Those with autoimmune disorders, however, viewed COVID-19 more as a personal threat, to be avoided with fewer trips outside the home. While those with respiratory disease were more likely to be fully vaccinated against COVID-19 than healthy controls, those with autoimmune disorders were not. When we controlled for disease status and response to survey 1, we found that behavior and attitude change was not dependent on having a chronic disease.

### Limitations

Adherence to or support for NPIs are self-reported. Social pressure on respondents to report greater adherence or support may have influenced responses at the time of sample. We note however that the measurement of relative difference between groups was highly significant across a variety of NPIs. It is difficult to imagine why these pressures would differ between groups; or why social pressures would be greater on individuals reporting chronic diseases than those without to the point that they confound otherwise insignificant results.

Representativeness of our sample may be limited as we utilized a convenience sample using quotas based on reported disease state. It would be extremely difficult to recruit sufficient chronic disease populations while maintaining representative sampling, however. This represents a necessary trade-off between our ability to test our key hypotheses about chronic respiratory and autoimmune diseases, and our ability to make statements about the general population. Samples also reflect the real-world social dimensions of these diseases, such the higher prevalence of these diseases among women.

Finally, when we compared the demographics of participants in survey 2 and survey 1 who did not participant in the resample, individuals with autoimmune diseases, non-Hispanic whites, women, people in the south, older respondents, and Republicans were more likely to participate in the second survey, while we were less likely to recontact students and Asian-Americans. Our analysis controls for these factors, however, and so the slight imbalance is unlikely to have impacted our results significantly.

## Conclusion

This research provides insight into how vulnerable individuals conceive of COVID-19 risk and adjust their behavior based on their disease status, which has implications for patient care and public health in general. The relationship between disease and acceptance of NPIs can shape how practitioners build support with individuals and communities for social and personal pandemic interventions. In maintaining public health measures in the long term for COVID-19 and other infectious disease outbreaks, individuals with chronic illnesses are likely to be more receptive and enduring supporters of public health interventions. Understanding the relationship between disease and vaccine acceptance allows us to address concerns of specific subpopulations to further promote vaccination against COVID-19.

## Data Availability

All deidentified data is archived by the authors and is available on request

## Abbreviations

ACS: American Community Survey
CI: Confidence Interval
NPI: Nonpharmaceutical intervention
OLS: Ordinary least squares

## Declarations

### Ethics approval and consent to participate

This study was approved by the US Naval Academy institutional review board, and informed consent was secured for all participants at the commencement of each survey.

### Consent for publication

Not applicable

### Availability of data and materials

The datasets used and/or analysed during the current study are available from the corresponding author on reasonable request.

### Authors’ contributions

BAS, EER, and JLK initially conceived of the study; all authors contributed to its design. NGE secured funding for the research. BAS conducted and monitored data collection. EER and BAS conducted data analysis. NGE wrote the first draft of the manuscript; all authors contributed to editing and final submission.

## Acknowledgements

*Not applicable*

