## Supplement for "COVID-19 risk perception and vaccine acceptance in individuals with chronic disease"

Supplementary Material

Appendix A: Full survey materials

COVID-19 is a serious threat to my health.

o Strongly agree

o Somewhat agree

o Neither agree nor disagree

o Somewhat disagree

o Strongly disagree

COVID-19 is a serious threat to the health of the public.

[Same response scale]

The threat of COVID-19 is overblown.

[Same response scale]

The coronavirus epidemic has changed some people's lives a lot, and other people's not at all. Please read each of the following statements and rate how true or false they are for you.

You are washing your hands more often and/or for a longer amount of time

o Very false for me

o Somewhat false for me

o Slightly false for me

o Slightly true for me

o Somewhat true for me

o Very true for me

You are working from home

[Same response scale]

You keep a distance of six feet between yourself and other people outside your home

[Same response scale]

You have reduced your trips outside of home

[Same response scale]

You wear a mask or other protection over your nose and mouth when you are outside your home

[Same response scale]

How much do you approve of the following ways to limit the spread of the COVID-19 pandemic?

Mandatory mask-wearing in public places

o Strongly approve

o Somewhat approve

o Slightly approve

o Slightly disapprove

o Somewhat disapprove

o Strongly approve

Limits on in-person worship or church services

[Same response scale]

Limits on in-door dining

[Same response scale]

Lockdown of all non-essential travel outside the home

[Same response scale]

Closure of schools

[Same response scale]

When a COVID-19 vaccine is made available to you, do you intend to get the vaccine?

o Definitely will get vaccine

o Probably will get vaccine

o May get vaccine

o May not get vaccine

o Probably will not get vaccine

o Definitely will not get vaccine

o I have already received a COVID-19 vaccine

[IF Definitely will/probably will/may get vaccine OR I have already received a COVID-19 vaccine]

What are the most important factors in your vaccine decision? Select all that apply.

▢ Want to protect myself from getting COVID-19

▢ Want to protect others from getting COVID-19

▢ Trust that the vaccine has been fully tested

▢ Trust that the vaccine is safe

▢ Want life to go back to normal

▢ Need the vaccine for work or other activities

▢ Other  _____________

[IF Definitely will/probably will/may not get vaccine]

What are the most important factors in your vaccine decision? Select all that apply.

▢ COVID-19 is not a serious health threat

▢ May have a bad reaction to the vaccine

▢ Feel that the vaccine has not been fully tested

▢ Feel that the vaccine is not safe

▢ Do not believe in vaccination

▢ Other  _____________

Do you identify as having a chronic illness, chronic disease, or disability? Check any that apply.

▢ Chronic illness

▢ Chronic disease

▢ Disability

▢ None of the above

Do you currently have any of the following chronic health conditions? Check any that apply. Otherwise, just skip this question.

▢ Cancer

▢ Asthma, Chronic Obstructive Pulmonary Disease (COPD), or any other chronic respiratory condition

▢ Any auto-immune disorder

▢ Diabetes

▢ Any cardiovascular disease

▢ Other   ________________________________________________

Have you ever tested positive for COVID-19?

o Yes

o No

Do you have any family members or close friends who have ever tested positive for COVID-19?

[Same response scale]

Generally speaking, do you usually think of yourself as a Republican, a Democrat, an Independent, or what?

o Democrat

o Republican

o Independent

o Other party

[IF Republican or Democrat]

Would you call yourself a strong [Republican/Democrat] or a not very strong [Republican/Democrat]?

o Strong [Republican/Democrat]

o Not very strong [Republican/Democrat]

[IF Independent]

Do you think of yourself as closer to the Republican Party or the Democratic Party?

o Yes, Democratic

o Yes, Republican

o No, Neither

Finally, we would like you to give us a little information about yourself. Please respond to the following background questions.

What is your age?

[Open numeric entry]

What is your gender?

o Male

o Female

o Other   ________________________________________________

Which of the following ethnicities describes you? Select all that apply.

▢ Latino/Hispanic

▢ Black/African-American

▢ Asian/Asian-American

▢ White/Caucasian

▢ Native American

▢ Other   ________________________________________________

What is the highest level of education that you have completed?

o Less than High School

o High School/GED

o Some College

o 2-year College Degree

o 4-year College Degree

o Master’s Degree

o Doctoral Degree (Ph.D., J.D., M.D., etc.)

What is your total family income (including parent income if they list you as a dependent)?

o Less than $10,000

o $10,000-$19,999

o $20,000-$29,999

o $30,000-$39,999

o $40,000-$49,999

o $50,000-$59,999

o $60,000-$69,999

o $70,000-$79,999

o $80,000-$89,999

o $90,000-$99,999

o $100,000 or greater

 In which state do you currently reside?

[Drop-down of US States and territories]

Use the scale below to indicate what kind of place you live in most of the time, where 0 is very urban, 2 is suburban, 4 is very rural, and the other numbers are in between. Please note you must move or click the slider to indicate a response.

[5-point slider scale]

[Student and employment status derived from Prolific stored data
